# Dynamics and future of SARS-CoV-2 in the human host

**DOI:** 10.1101/2020.07.14.20153270

**Authors:** Michael Gillman, Nuno Crokidakis

## Abstract

Forecasting trends in COVID-19 infections is vital for the global economy, national governments and physical and mental well-being. Using the *per capita* number of new cases as a proxy for the abundance of the SARS-CoV-2 virus, and the number of deaths as a measure of virulence, the dynamics of the pandemic and the outcomes emerging from it are examined for three locations (England, Italy and New York State). The data are analysed with a new version of a population dynamics model that combines exponential/logistic growth with time-varying carrying capacity, allowing predictions of persistence or extinction of the virus. In agreement with coevolutionary theory, the model suggests a transition from exponential virus growth to low abundance, coupled with reduced virulence, during colonisation of the alternate human host. The structure of the model allows a straightforward assessment of key parameters, which can be contrasted with standard epidemiological models and interpreted with respect to ecological and evolutionary processes and isolation policies.

## Introduction

There are many possible options for the future of COVID-19, its effects on social and political structures and the consequences for the wider environment; in short, the transition to, and the form of, the ‘new normal’. The coevolution of hosts and viruses provides an important context for the dynamics and possible futures of the coronavirus SARS-CoV-2 in humans. Large shifts in incidence and virulence of viruses occur between natural hosts, which may experience high constant levels of infection but low symptoms, and alternate hosts of a different species (Vijaykrishna et al. 2007). Colonisation of alternate hosts may be associated with exponential growth of the virus population and high levels of symptoms. Following this, either extinction of the virus in the alternate host or an evolutionary shift towards coexistence in a less virulent state is expected (Ye et al. 2020).

Of the seven known human coronaviruses (including SARS-CoV-2), the four endemic species are considered benign viral pathogens for humans (Li 2013) causing ‘mild, self-limiting respiratory infections’ (Fehr and Perlman 2015). Neither of the previous two more virulent species (MERS-CoV and SARS-CoV) have persisted in the alternate human hosts (MERS-CoV-2 has sporadic outbreaks associated with infected camels but no known human to human transmission, WHO 2019). In contrast, the four less virulent species circulate continuously (Drexler et al. 2014) with frequencies of up to a few percent in patients with respiratory tract disease (Sloots et al. 2006, Esper et al. 2006, Gaunt et al. 2010). Five of the seven human coronaviruses are believed to have originated in bats (HCov-229E, MERS-CoV, SARS-CoV, SARS-CoV-2 and HCoV-NL63, Lau et al. 2005, Li et al. 2005, Huynh et al. 2012, Drexler et al. 2014, Ye et al. 2020, Zhou et al. 2020) with the other two having rodents as their primary host (HCoV-OC43 and HCoV-HKU1, Ye et al. 2020).

While the absolute abundance of SARS-CoV-2 or other viruses in their hosts are difficult to estimate, assessing the changes in relative abundance, temporally or spatially, is achievable using different proxies. Population dynamics models of COVID-19 infections have included developments of classic epidemiological and ecological models; from standard Susceptible-Infected-Recovered (SIR) to models with quarantine, Crokidakis 2020, to eight compartment models, Giodrano et al. 2020, and logistic to generalised Richards model, Roosa et al. 2020. Modelling efforts have been motivated by the need to understand the effects of social distancing or containment policies in the absence of vaccines and antivirals (Chowdhury et al. 2020, Flaxman et al. 2020, Maier and Brockmann 2020). Forecasting models have predicted persistence of SARS-CoV-2 for several years with assumptions of seasonal variation and cross-immunity with endemic human coronaviruses (Kissler et al. 2020). Here we use a novel population dynamics model to quantify changes in the incidence of SARS-CoV-2 during the pandemic and address the hypothesis that the SARS-CoV-2 virus in its new human host will either tend towards extinction or low densities with reduced virulence.

## Methods

We test the hypothesis with a regression model of virus dynamics, contrasting new cases, as a measure of virus abundance, with deaths, as a measure of virulence. The temporal dynamics of new COVID-19 cases is not only a measure of the change in numbers of infected people with time, but also changes in the incidence of the virus causing those infections. Summing the number of new cases over consecutive days provides an estimate of the total virus incidence or abundance (hereafter the term abundance is used), assuming a dynamic compartment model in which hosts enter from a susceptible, or exposed, state through an infected state to a recovered (or deceased) state. The incubation period (time from exposure to symptom onset) has been estimated to be between 4.5 and 5.8 days for 95% of cases (median 5.1, Lauer et al. 2020) with 97.5% people developing symptoms within 11.5 days. Here we use a thirteen-day window to encompass the life cycle of the virus in most human hosts and for smoothing weekly reporting. This also agrees with the five-day incubation period plus eight-day (long) delay from onset to isolation used in Hellewell et al. (2020).

Expression of virus abundance as a fraction of the host abundance facilitates more meaningful comparisons between locations and allows insights into population regulation and therefore persistence or extinction of the virus. The new case data are reported and analysed as thirteen-day totals per million people (referred to as ncpm). Dates are referenced to the midpoints of the thirteen-day totals. All dates are for the year 2020 unless otherwise indicated. The first value of the first thirteen-day total (*N*_0_) is set at the date at which the cumulative number of cases in a location exceeded 100 (2 March England, 23 February Italy and 9 March New York State). The new case data are contrasted with fatality data using the same variable of thirteen-day totals per million people (fpm), noting that the date of death may be highly variable with respect to the date of infection. The first day of the thirteen-day death totals is the first recorded virus-associated death in the location (7 March England, 21 February Italy and 14 March New York State). The subset of new cases which end in fatalities is partly a reflection of the age and health profile of those infected, for example, 90.0% of 24,959 fatalities (correct at end of 9 July 2020) in New York State occurred in people with one or more comorbidities such as hypertension and diabetes (New York State Department of Health 2020).

A simple model of population dynamics is provided by the logistic equation, which can be described in continuous or discrete time forms producing characteristic s-shaped dynamics of abundance (*N*) over time, with options for more complex dynamics in the discrete form (May and Oster 1976).

The continuous logistic equation

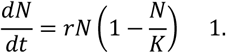

combines *r*, the maximum rate of population growth, with *K*, the carrying capacity, which mimics a limiting resource such as the number of susceptible individuals. In ecological studies of population growth, within a single species, the reduction effect of functions like 1-(*N*/*K*) on the rate of change in *N* is an example of a density-dependent process, with survival or fecundity changing with increasing density (Gillman 2009). The result is regulation of the size of *N*, which eventually reaches a steady-state value (equilibrium) of *K*. As new cases are a proxy for virus abundance the resulting parameter estimates are comparable with standard ecological studies of population growth.

Fitting a logistic model to the changing numbers of *new* cases is rare (e.g. Shen 2020) with most published models of COVID-19 infection using the logistic or similar equations to fit the *cumulative* numbers of cases or deaths (e.g., Almeshal et al. 2020, Batista 2020, Gaeta 2020, Jia et al. 2020, Kamrujjaman et al. 2020, Roosa et al. 2020, Tátrai and Várallyay 2020, Vasconcelos et al. 2020). Having fitted the logistic or a more complex version (e.g. the Generalised Richards Model, Biswas and Roy 2020, Roosa et al. 2020, Vasconcelos et al. 2020; five parameter model, Kriston 2020) to cumulative cases, it is then possible to reconstruct the change in new cases through time. The relationship between the Richards modification of the logistic model and SIR models is discussed in Wang et al. (2012).

With studies of cumulative cases, *K* indicates the final (total) number of cases. In so doing it assumes that the number of new cases will eventually reach zero. In contrast, *K* within a model of new cases as a measure of virus abundance is indicative of the ongoing carrying capacity or steady state of the virus in a given host population. The assumption of a steady state value for new cases is only reasonable if new host resources (susceptible individuals) are continually available. Therefore, a straightforward modification to the model is to assume that there is not a continual supply of susceptible individuals, i.e., *K* changes with time. Reductions in *K* are expected due to a combination of natural processes (immunity, death) and legislative measures which may enhance natural processes (social distancing acting to mimic avoidance of infected individuals, e.g., Kiesecker et al. 1999). Increases in *K* may be artefacts of sampling but can also incorporate dispersal across heterogeneous host landscapes.

The model of virus abundance with time (*N*_*t*_) employed here combines logistic growth with a time-dependent carrying capacity (*K*_*t*_). Continuous time models with the logistic equation and time-varying carrying capacity are mathematically difficult, with few exact solutions (Shepherd and Stojkov 2007, Lanteri et al. 2020). Therefore, we use a discrete-time analogue (difference equation) in which λ is the maximum rate of population growth (ln (λ) is equivalent to *r* in Equation 1):

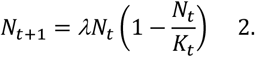

There are various options for the function *K*_*t*_ (Coleman 1978, Banks 1994, Cohen 1995, Meyer and Ausubel 1999, Safuan et al. 2013). Here we use:

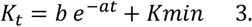

which has the value *b* + *Kmin* at time 0 and tends towards *Kmin* with time (*t*), with options for long-term persistence (*Kmin*>0) or extinction (*Kmin*<0). Negative values of *Kmin* are not biologically meaningful but give an indication of the likelihood of reaching zero and therefore extinction of the virus. The population equilibrium value (*N*_*e*_) for the discrete logistic with constant *K* is *K* (1-1/λ). With the function *K*_*t*_ in Equation 3, the model will therefore tend towards a *N*_*e*_ value of *Kmin* (1-1/λ) with time. The exponential decay also mimics change in susceptibles in standard SIR compartment models with parameter *a* determining the rate of change with time. The time steps are one day, therefore N_t+1_/N_t_ approximates continuous change in virus abundance. Application of the logistic and time-dependent carrying capacity model to the number of fatalities follows a similar logic to new cases in that it is a model of the change in virus abundance but with a particular outcome for the host.

Using nonlinear regression (nls, R core team 2018) the parameter values can be extracted from a time-series and then used in a reconstruction of the dynamics with extrapolation as appropriate. Parameter estimation was checked with a model time-series in which the four parameters were set (with N_0_ = 1) and all were estimated to very high accuracy.

The hypothesis is tested here with data from England, Italy and New York State which provide sufficiently robust data to evaluate the method and illustrate possible outcomes (data correct at 12:00 GMT on 13 July 2020 and sources listed in References under location name). Exploration of the data and residuals from model fits suggested that some of the new case data were too low in the peak period of April. This was especially apparent when contrasting the change in the England data after wider population data started to be included and is supported by compartment models of Italian cases (Pedersen and Meneghini 2020). To avoid these issues, and illustrate the potential under-reporting during the peak period, the case models are fitted to the ascending and descending sections of the data (omitting 1 to 27 April for England, 16 March to 22 April for Italy and 28 March to 7 April for New York State; omissions judged with respect to R^2^). The full sets of available data are fitted for the fatality models.

## Results

The model fits for new cases and deaths reveal a tight range of peaks within 20 to 33 days of the start (as defined in the methods), followed by a slow descent over 100+ days (Figure 1). The model peaks for new cases, predicted from data either side of the peak, are up to twice the value of the observed (Italy model 2411 and observed 1161 ncpm). The peak model cases and deaths for New York State (7860 ncpm, 657 fpm) are over three times the values for England and Italy (England 1239 ncpm and 206 fpm; Italy 2411 ncpm and 186 fpm).

**Figure 1.**
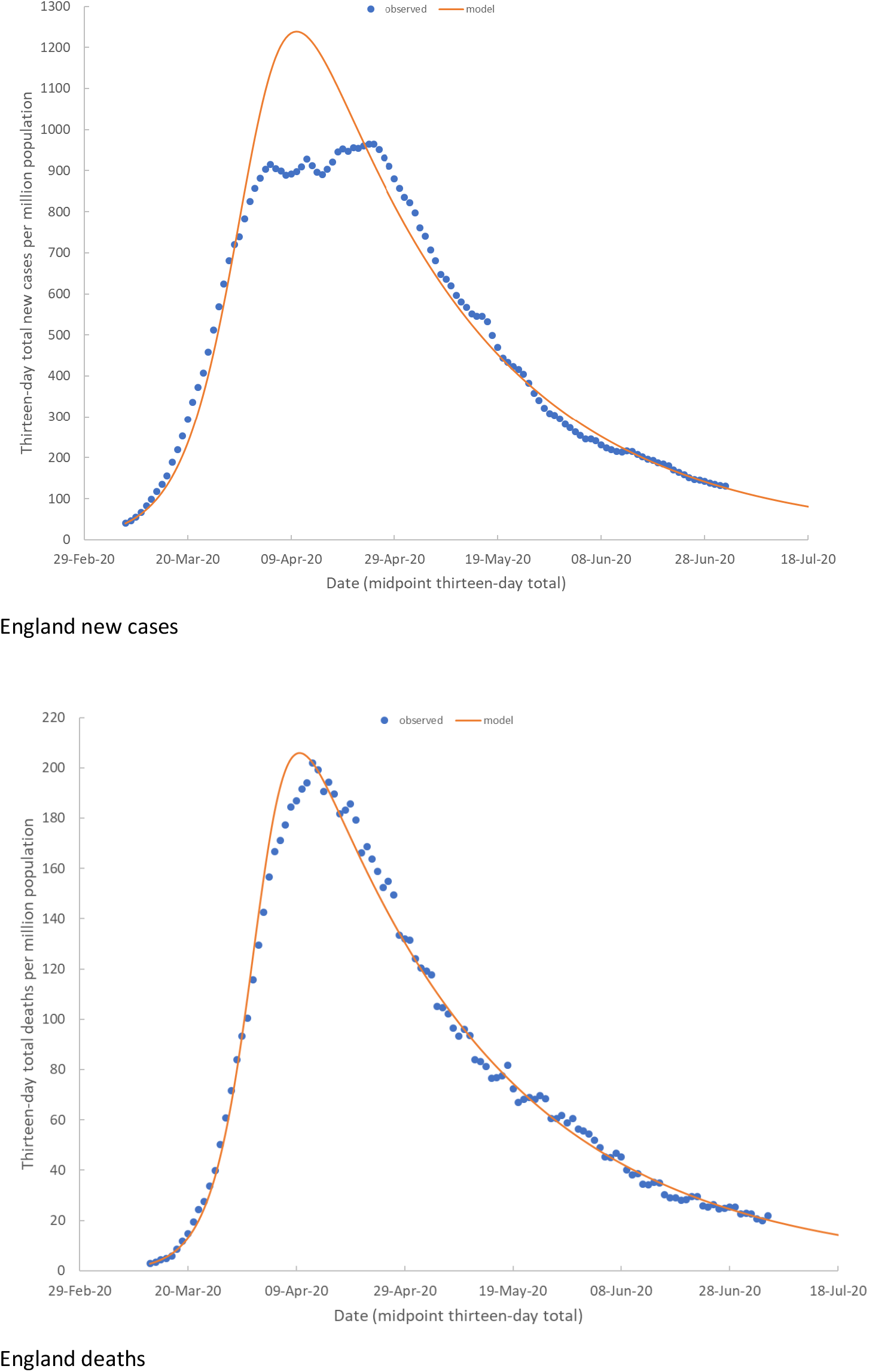
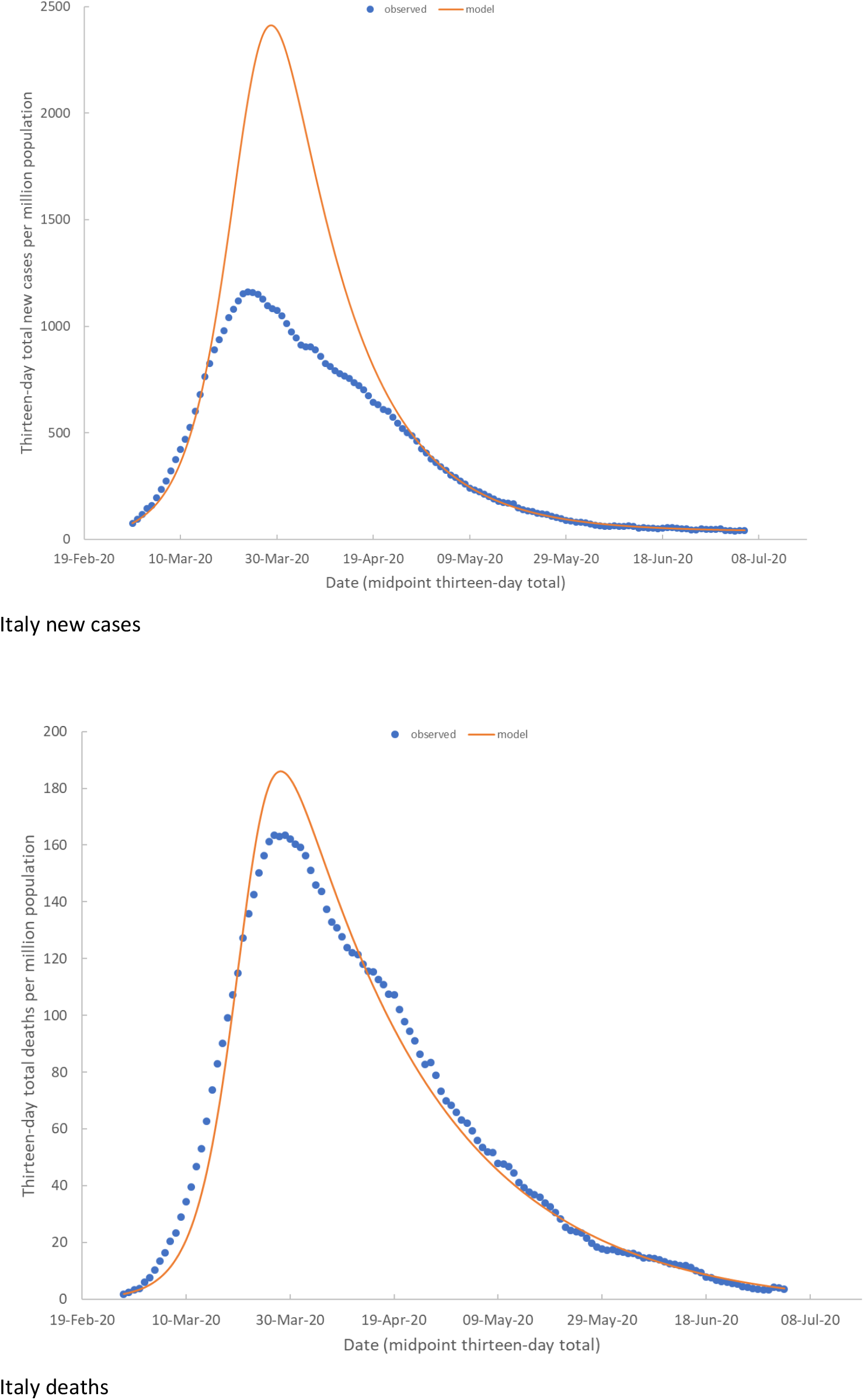
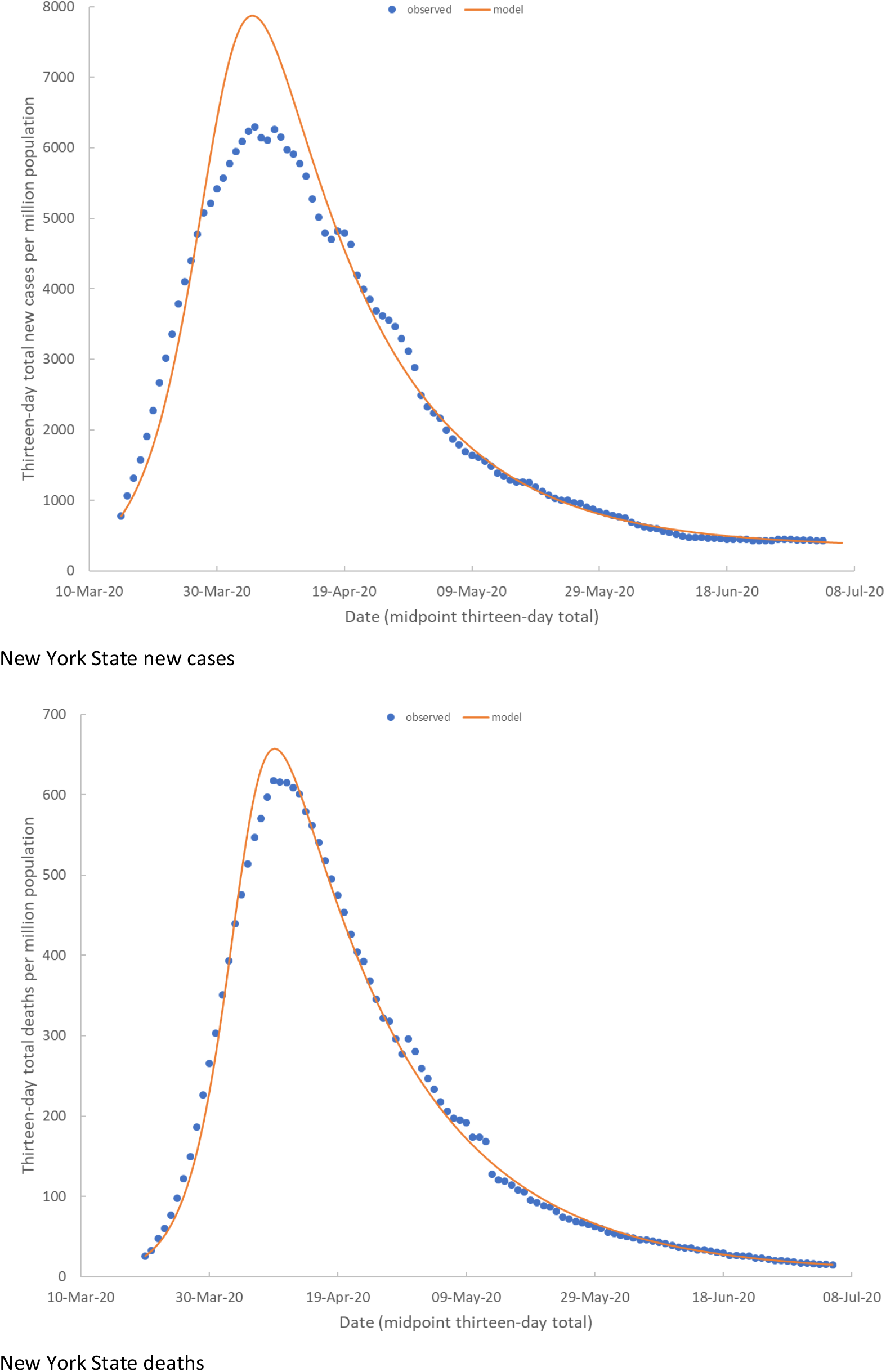
Observed values and model fits. New cases and deaths for three locations.

The rate of virus population growth, λ, is higher for deaths than new cases (averages of 1.25 and 1.18; Table 1, Figure 2). The *K*_*t*_ shape parameter (*a*) falls into two groups with averages of 0.03 and 0.06 (Figure 2). Population equilibrium values for deaths are low to negative (5.7 to -3.1 fpm) with greater variation in the three positive values for new cases from 6.6 for England to 340 ncpm for New York State (Table 1).

**Table 1.**
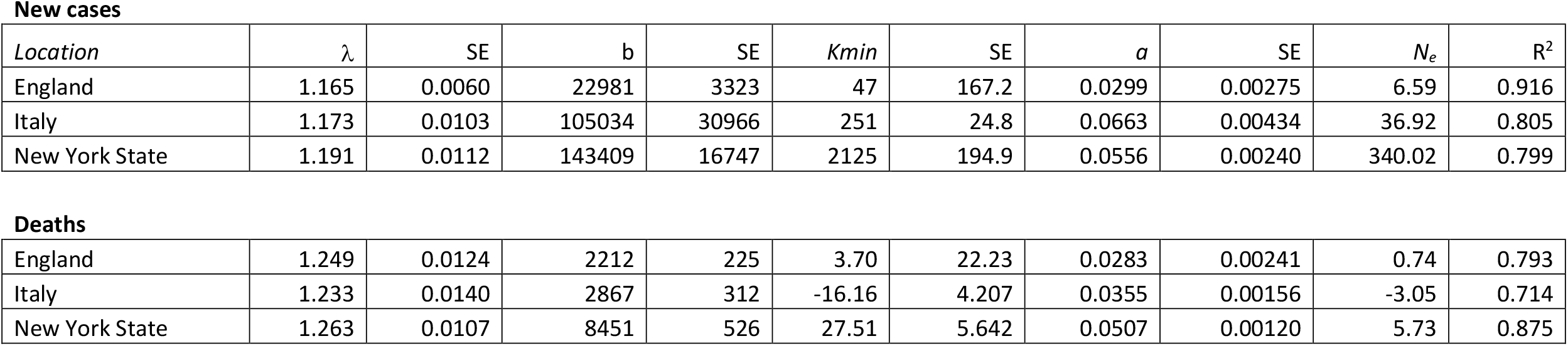
Parameter values and standard errors (SE) from the discrete logistic model with time-varying carrying capacity.

**New cases**
| *Location* | $\lambda$ | SE | b | SE | *Kmin* | SE | *a* | SE | $N_e$ | $R^2$ |
| --- | --- | --- | --- | --- | --- | --- | --- | --- | --- | --- |
| England | 1.165 | 0.0060 | 22981 | 3323 | 47 | 167.2 | 0.0299 | 0.00275 | 6.59 | 0.916 |
| Italy | 1.173 | 0.0103 | 105034 | 30966 | 251 | 24.8 | 0.0663 | 0.00434 | 36.92 | 0.805 |
| New York State | 1.191 | 0.0112 | 143409 | 16747 | 2125 | 194.9 | 0.0556 | 0.00240 | 340.02 | 0.799 |
| **Deaths** |  |  |  |  |  |  |  |  |  |  |
| England | 1.249 | 0.0124 | 2212 | 225 | 3.70 | 22.23 | 0.0283 | 0.00241 | 0.74 | 0.793 |
| Italy | 1.233 | 0.0140 | 2867 | 312 | -16.16 | 4.207 | 0.0355 | 0.00156 | -3.05 | 0.714 |
| New York State | 1.263 | 0.0107 | 8451 | 526 | 27.51 | 5.642 | 0.0507 | 0.00120 | 5.73 | 0.875 |

**Figure 2.**
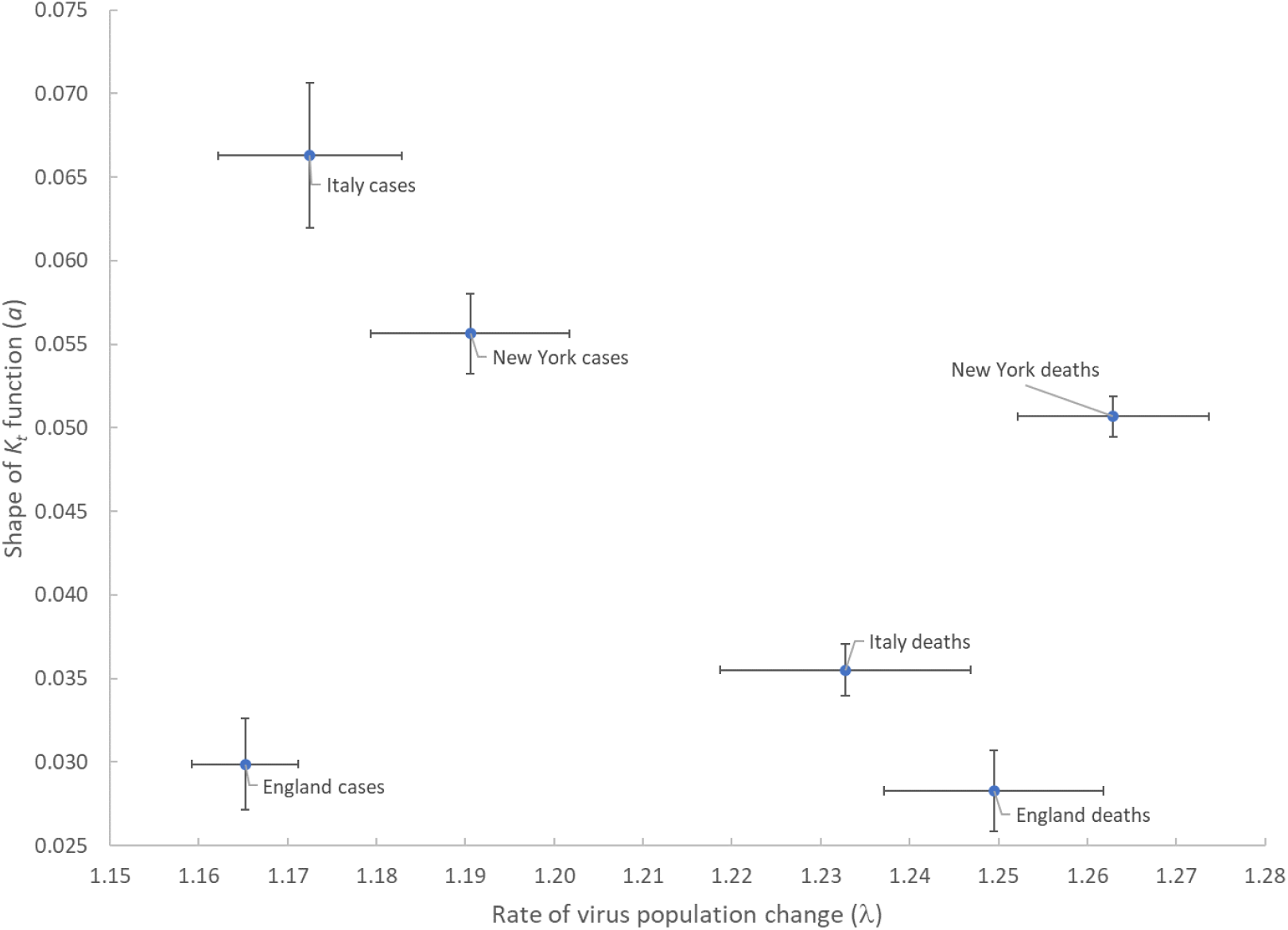
Model parameters *a* versus λ and standard errors.

## Discussion

None of the three locations are currently predicted to have negative equilibrium values for new cases, suggesting persistence of the virus (although the England *Kmin* value is not significantly different from zero). The death equilibrium values are either negative (Italy) or less than 1 in 10,000. An important assumption is that these equilibrium values continue under relaxed isolation conditions, i.e., they are stable with respect to future social behaviours. Interpretation of the size of the equilibrium population values for new cases also depends on the adequacy of sampling. If the New York equilibrium value is taken as an upper value, then about 0.03% of people would be expected to be infected at any one time. The random household studies of SARS-CoV-2 by the Office for National Statistics (ONS 2020) in England gives values 1.7 to 3.7 times higher than the thirteen-day incidence used here (five sample dates matched). Therefore, the equilibrium value might be close to 0.1% for New York State. Sweden, which did not impose a strict lockdown but implemented some restrictions such as closure of secondary schools, provides an interesting contrast to the data analysed here (Sweden data 10 July). Their new cases were consistent at around 700 ncpm for about a month before rising to 1400 and now dropping (less than 1000 on 10 July). Conversely, deaths have shown a consistent linear reduction from a peak in mid-April (122 fpm) to less than 20 fpm by the end of June. These data therefore also suggest a maximum persistence of around 0.1% with a negligible contribution to mortality.

In addition to predicting future possible outcomes, the model is useful for assessing potential abundance during the pandemic. Notably, the fit to either side of the peak for new cases suggests an up to two-fold underestimate in virus incidence during the peak. One test of the model is to contrast parameter estimates with those of epidemiological models. The reproduction number *R*_0_ in the early stages of the COVID-19 outbreak has been estimated as 2.38 (Li et al. 2020) with transmission models assuming 2.2 or similar (e.g., Chowdhury et al. 2020). With a generation time (serial interval) of 5 days (Ferretti et al. 2020 5 days ± 1.9 SD; Nishiura et al. 2020 4.6 days, 3.5 – 5.9 95% CI) an *R*_0_ of 2.2 equates to an *r* of 0.158 (ln(2.2)/5), which in turn equates to a λ of 1.17, in agreement with the model estimates for England and Italy. The exponential decay in *K*_*t*_ is governed by the parameter *a* which can be related to the depletion of susceptibles and explained by isolation policies (Crokidakis 2020, Maier and Brockmann 2020). A higher value of *a* can be interpreted as indicating more effective isolation policies, alongside natural processes such as immunity (note that the carrying capacity function assumes reduction from *t*=0 prior to implementation of lockdowns in the examples used here), including cross-immunity with other coronaviruses (Petrosillo 2020). The Italy and New York State new cases have values of *a* which are approximately twice that of England new cases, perhaps suggesting more effective control measures.

An important attribute of the model is that it can easily be applied to other epidemics. An obvious comparison is with the SARS epidemic in 2003. Using five-day averages of new case data presented in Hung 2003 (his Phase 2 and first half of Phase 3 from 2 April to 20 May 2003) the model produces estimates for λ of 1.22 ± 0.059, 0.055 ± 0.013 for *a* and -2.65 ± 18.7 for *Kmin* with an *R*^2^ of 42.7%. The estimate of λ is between the SARS-CoV-2 values for cases and deaths with *a* in the higher group of SARS-CoV-2 estimates. Extrapolating forward with this model gives an extinction date of 2 July 2003 and extrapolating back gives a start date (1 case over 5 days) of 7 March 2003. The predicted start date is two weeks after the first (index) case on 21 February 2003 (SARS 2003). The extinction point is three weeks after the last probable date on 11 June 2003 (WHO 2003). Therefore, the model prediction of 117 days is in good agreement with the observed 110 days.

## Conclusion

The discrete logistic with time-varying carrying capacity provides a tractable model of changes in abundance of SARS-CoV-2 during the current pandemic. The predicted outcomes (equilibrium population values) suggest persistence of the virus with low virulence in agreement with coevolutionary theory. Parameter estimates are consistent across locations and easily detectable with time series, suggesting the wider application of the model to this and other infectious diseases. The extent to which this is a global phenomenon, and the relative contributions of social distancing and intrinsic dynamics of the virus in the alternate human host, merits further consideration.

## Data Availability

Data are in the public domain and cited in references

## Acknowledgements

Our thanks to Hilary Erenler for detailed comments. NC acknowledges financial support from the Brazilian funding agencies CNPq and FAPERJ.

